# Software Application Profile: A real-time surveillance system for monitoring heat exposure and its health impacts - presenting the Rio de Janeiro Heat Dashboard

**DOI:** 10.64898/2026.08.26.26361449

**Authors:** João Henrique de Araujo Morais, Caroline Dias Ferreira, Valeria Saraceni, Débora Medeiros de Oliveira e Cruz, Gislani Mateus Oliveira Aguilar, Oswaldo Gonçalves Cruz

## Abstract

**Motivation:** With the scaling frequency and intensity of extreme heat events across the globe, it is critical for public institutions to develop early detection systems and continuous monitoring of these events and their impacts. In Brazil, Rio de Janeiro was the first city to publish its heat protocol, with the Rio Heat Dashboard as a central component of this system.

**Implementation:** The dashboard was implemented using R/Shiny and integrates climatic and health data from multiple sources.

**General features:** The application comprises real-time heat exposure monitoring and automatic alert level classification, which is monitored daily by multiple municipal actors and supports activation of actions specified in the heat protocol. It also features a health impact module, which lists each heat event and its impact on mortality, and primary care and emergency visits.

**Availability:** The source for full reproducibility is available through https://github.com/joaohmorais/RioHeatDashboard.

## Introduction

Heat waves and extreme heat events represent scaling threats, especially in a climate change scenario. The frequency and intensity of such episodes are significantly increasing^1,2^, representing a significant toll on morbidity, mortality and healthcare costs^3^. Over the last decades, countries and public organizations have implemented Heat-Health Warning Systems (HHWS), which combine meteorological forecasts and public health actions in order to prepare for such events and reduce health impacts ^4^. For a HHWS to effectively work, cities require systems that provide timely weather information to generate alerts and guide response actions. Initiatives like the HEAT-SHIELD platform ^5^, which provides forecast and custom worker safety recommendations, and forecaster.health (https://forecaster.health/), that produces temperature warnings based on mortality estimates, can be named as functional examples.

While HHWS exist and are formally described in Europe and North America ^6^, Latin American countries have lagged dramatically in the implementation of such systems, and Rio de Janeiro was the first Brazilian city to publish a formal Heat Protocol in 2024 ^7^. Rio’s approach differs from others by considering exposure period as one of its trigger metrics, and for having a near-real time health impact monitoring embedded in its Heat Monitoring Dashboard (*Painel de Calor*). In this Software Application Profile, we describe the modules and functionalities of the Heat Monitoring Dashboard (*Painel de Calor*), a platform conceived and developed by the Epidemiological Intelligence Center of the Rio de Janeiro Municipal Health Secretariat. Special attention is given to its replicability potential and to the integration of global weather forecasts, meteorological station measurements, and health data in order to estimate health impacts in near real time. The platform also operates as an early warning system for triggering the Heat Protocol of the City of Rio de Janeiro, supporting timely risk assessment, decision-making, and coordinated public health response during extreme heat events.

### Implementation

The *Painel de Calor* R/Shiny app was developed using the {golem} framework, which allows users to easily install and run it like an R package in various environments. The app’s source code and example data are hosted in the GitHub repository https://github.com/joaohmorais/RioHeatDashboard. This section describes the automatic data ingestion process and the role of each module of the application.

#### Data ingestion

Weather station hourly measurements of temperature (T) and relative humidity (H) are collected from three distinct sources: INMET (national weather service), REDEMET (airports and airforce bases), and Alerta Rio (municipality stations). The heat index (HI) is then calculated based on NOAA’s operationalization (https://www.wpc.ncep.noaa.gov/html/heatindex_equation.shtml). An automated R script is executed every fifteen minutes and gathers any new measurements from each source, consolidating the observations in a single database implemented in DuckDB.

Global prediction models forecasts are retrieved three times a day from the OpenMeteo service (https://open-meteo.com/) via its open API. Hourly forecasts of climatic variables are retrieved for up to 7 days in advance. Currently considered models for Rio are (provider, resolution): ICON (DWD/Germany, 11km), IFS (ECMWF/Europe, 9km), GFS (NOAA/United States, 13km), GEM (Canadian Meteorological Service/Canada, 15m), GSM (JMA/Japan, 55km) and ARPEGE (Météo-France/France, 25km).

Electronic health records from Rio municipality’s primary care and emergency units, as well as mortality registries, constitute the health data also inputted to the app. This information is integrated into a DuckDB database via a nightly Extract, Treat and Load (ETL) process, which gathers data from all visits in each unit from the previous day. Mortality registries are updated weekly.

#### Daily monitoring and alert classification

The main screen of the application presents a daily view, showing the hourly measurements for T and HI, as well as the forecasts in a single visualization (Figure 1). The values shown represent the hourly median across all the stations (A), but individual measurements can be seen as well (B). Summary measures are calculated and presented on the right-hand side, such as the highest HI value observed (C), and the amount of hours spent above each heat stress threshold (D). The latter, along with the weekly context (E) are used to calculate the suggested Heat Level for the city (F), according to the logic defined in the protocol (ref.).

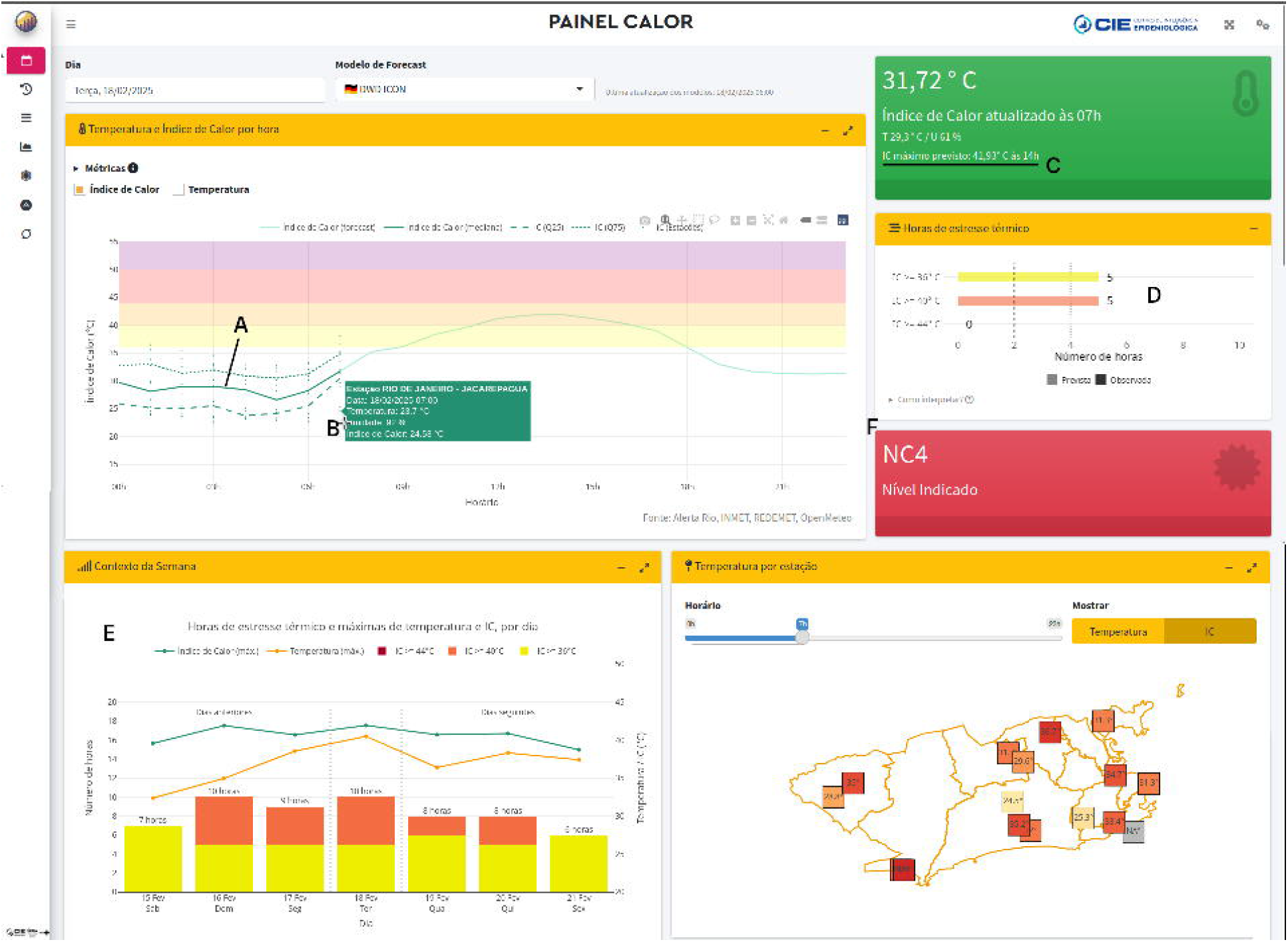

#### Model validation module

The second tab for the application provides an evaluation of the performance by each of the included global prediction models. The forecasted and observed HI curves can be compared visually, and Root Mean Squared Error (RMSE) metrics are interactively shown for a chosen period: last 1, 3, 7, 14 or 30 days.

#### Historical series module

This module consists of a calendar-based visualization interface that allows users to observe the classification of each day according to heat levels, alongside a summary table presenting the distribution of days by heat level category.

#### Health impact module

Finally, the health impact screen summarizes the direct impact of each heat event on different health outcomes: primary care visits (PCVs), emergency visits (EVs) and mortality. A heat event is classified as a sequence of days for which at least Heat Level (HL) 3 was observed uninterruptedly. For each event, the following metrics are shown: highest HL reached, daily maximum HI, event duration (days), and observed (O), expected (E) and O/E ratio for PCVs, EVs and mortality (Figure 2). For each day of the event, the expected value of visits or deaths is calculated as the median daily counts due to the considered causes in the preceding 365 days, in that same week day. The included causes for PCVs and EVs are (ICD-10 codes, description): T67 (Effects of heat and light), R55 (Syncope and collapse), I95 (Hypotension), E86 (Volume depletion), L55 (Sunburn), L74.0 (Miliaria rubra) and I21 (Acute myocardial infarction). For mortality, the user may select between natural or total deaths. For deaths, the day following the event end date is also included to account for part of the mortality displacement lag ^8^.

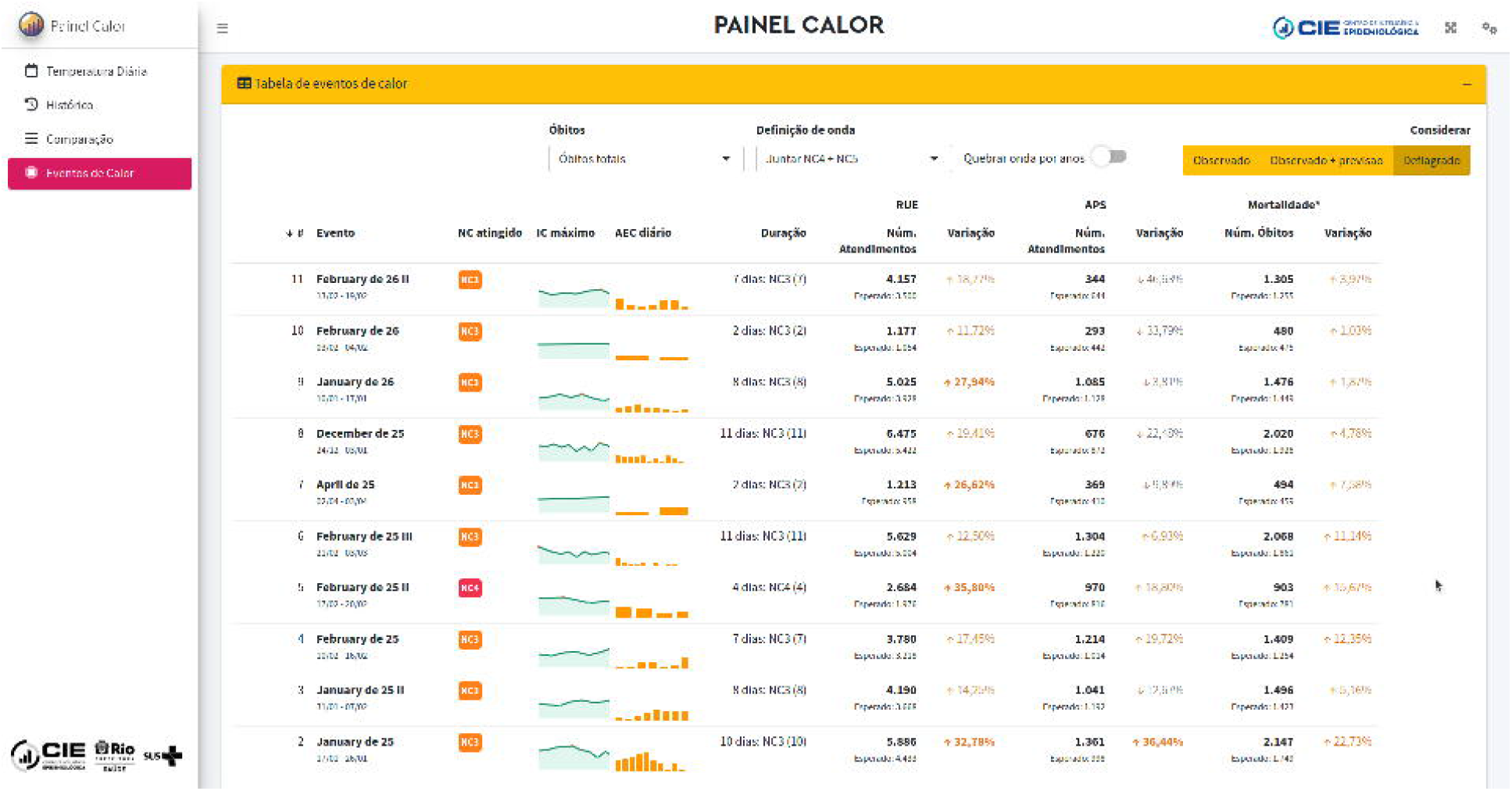

### Use

The Heat Dashboard is accessed by the following actors in the municipality: Health Surveillance department (SVS) of the Municipal Health Secretariat, Operations and Resilience Center (COR), Meteorological Alerts department (Alerta Rio), and the Secretariat for Environment and Climate (SMAC). The monitoring is done on a daily basis, even on weekends and holidays -since part of the mentioned actors include workforce on call 24 hours a day. The application is hosted in a virtual machine in COR infrastructure, being available for access in Rio’s municipality internal network.

Daily, Alerta Rio meteorologists monitor the heat conditions for the present day as well as for 3 days in advance. In case one or more global prediction models indicate the possibility of elevating the HL (automatically calculated and shown in the main dashboard tab - Figure 1, F) an internal communication is spread. If the prediction does confirm - a median value for HI above the defined thresholds is observed among the stations - another communication is sent indicating the triggers have been reached and suggesting the HL change. When that happens, communication measures are immediately taken, and for HL 4 and beyond adaptation or suspension of activities may be implemented.

We take for example the first occurrence of a HL 4. On January 15th, 2025, HL 3 was already active and models indicated the possibility of a HL 4 on Monday, 17th. The first internal communication procedures were implemented and on Sunday, 16th, Rio Mayor Eduardo Paes called a press conference to alert the population for the possible HL 4 (ref.), its impacts and actions from the City Hall, such as the opening of cooling zones (ref). On Monday, the triggers for HL 4 were reached indeed, which led to the first - and so far, only - occurrence of a HL 4 in the city, which lasted for 4 days (ref.).

Through the health impacts tab in the dashboard (Figure 2), it is possible to see that the HL 4 event led to an increase of 35,80% in emergency visits (the highest so far) and 16,30% in natural causes mortality. These metrics are further used in reports produced after each heat event in the municipality and to monitor healthcare system strain during periods of extreme heat.

## Discussion

The Heat Dashboard presented in this paper plays a crucial role in the implementation and maintenance of Rio de Janeiro’s Heat Protocol. Through this system, current heat conditions are continuously monitored, and alerts are generated whenever predefined thresholds are reached. The innovative integration of observations and forecasts from multiple global weather models into a single visualization, together with the automatic calculation of the suggested heat level for the current and upcoming days, enables preparedness, risk communication, and timely response in real time.

In addition, the health impact module, updated with near real-time healthcare attendance data (1-day lag), allows the health surveillance team to monitor and quantify the impacts of each heat event on population health while the event is ongoing. Integrated into the operational framework of the municipal Heat Protocol, the system functions both as an early warning system for extreme heat events and as a near real-time monitoring platform to support decision-making and guide response actions. The platform also supports retrospective analysis of heat events, enabling comparisons between different episodes and providing evidence to refine operational strategies, improve protocols, and strengthen future preparedness and response actions.

Important limitations in the whole data generation process need to be stated. Even though weather station measurements from three sources were joined, the stations’ locations are still uneven across Rio, and many densely populated regions are not represented. The choice of using the median across all the stations also may not capture when specific parts of the city are particularly affected. Also, the forecast products obtained are not optimized or downscaled for Rio, which results in particularly large pixels for the city and may smooth important patterns. Regarding the health impact module, since the expected and observed metrics are calculated in running time, they follow a relatively simple formulation: the day-of-week median for the last year. This way, no adjustment is made for other possible confounders such as possibly ongoing epidemics in the city, air pollution, or seasonality.

A strength of the developed app is its modularization and publicly code sharing. These allow for replication among other cities and contexts, once data sources are adequately adapted for data ingestion, and specific triggers are defined. It also keeps it relatively simple to add new data sources or functionalities. In the current moment, Rio has acquired 21 low-cost sensors, which provide temperature and humidity measurements in a finer spatial distribution across the city. Future developments include, therefore, the possibility of running local estimates of heat exposure using the stations and sensor data. Additionally, more detailed information can be explored for the health impacts module - such as most distribution of visits and deaths among age and sex groups, most affected causes and locations.

## Data Availability

This study used electronic health records data from the Rio de Janeiro municipality which are not publicly available. In the GitHub repository, however, simulated data based on the real distributions were made available for reproducibility.

https://github.com/joaohmorais/RioHeatDashboard

## Acknowledgments

We would like to thank Rio’s Municipal Health Secretariat (SMS), Operations and Resilience Center (COR), Alerta Rio, and Municipal Secretariat for Public Services and the Environment (SMAC) for collaborative work.

